# Model-free estimation of COVID-19 transmission dynamics from a complete outbreak

**DOI:** 10.1101/2020.07.21.20159335

**Authors:** A. James, M. Plank, S. Hendy, R. Binny, A. Lustig, N. Steyn

## Abstract

New Zealand had 1499 cases of COVID-19 before eliminating transmission of the virus. Extensive contract tracing during the outbreak has resulted in a dataset of epidemiologically linked cases. This data contains useful information about the transmission dynamics of the virus, its dependence on factors such as age, and its response to different control measures.

We use Monte-Carlo network construction techniques to provide an estimate of the number of secondary cases for every individual infected during the outbreak. We then apply standard statistical techniques to quantify differences between groups of individuals.

Children under 10 years old are significantly under-represented in the case data. Children infected fewer people on average and had a lower secondary attack rate in comparison to adults and the elderly. Imported cases infected fewer people on average and had a lower secondary attack rate than domestically acquired cases. Superspreading is a significant contributor to the epidemic dynamics, with 20% of cases among adults responsible for 65-85% of transmission. Asymptomatic cases infected fewer individuals than clinical cases. Serial intervals are approximately normally distributed (*μ* = 5.0 days, *σ =* 5.7 days). Early isolation and quarantine of cases reduced secondary transmission rates.

Border controls and strong social distancing measures, particularly when targeted at superspreading, play a significant role in reducing the spread of COVID-19.

## Introduction

Between 26 February and 22 May 2020, New Zealand recorded 1499 confirmed and probable cases of COVID-19 (Figure 1). Of these, 575 had a recent history of overseas travel. With the exception of international arrivals, which are subject to mandatory 14-day quarantine, no community cases were reported from 22 May to 10 August. This means that New Zealand has a high probability of having eliminated community transmission during this period.

**Figure 1:**
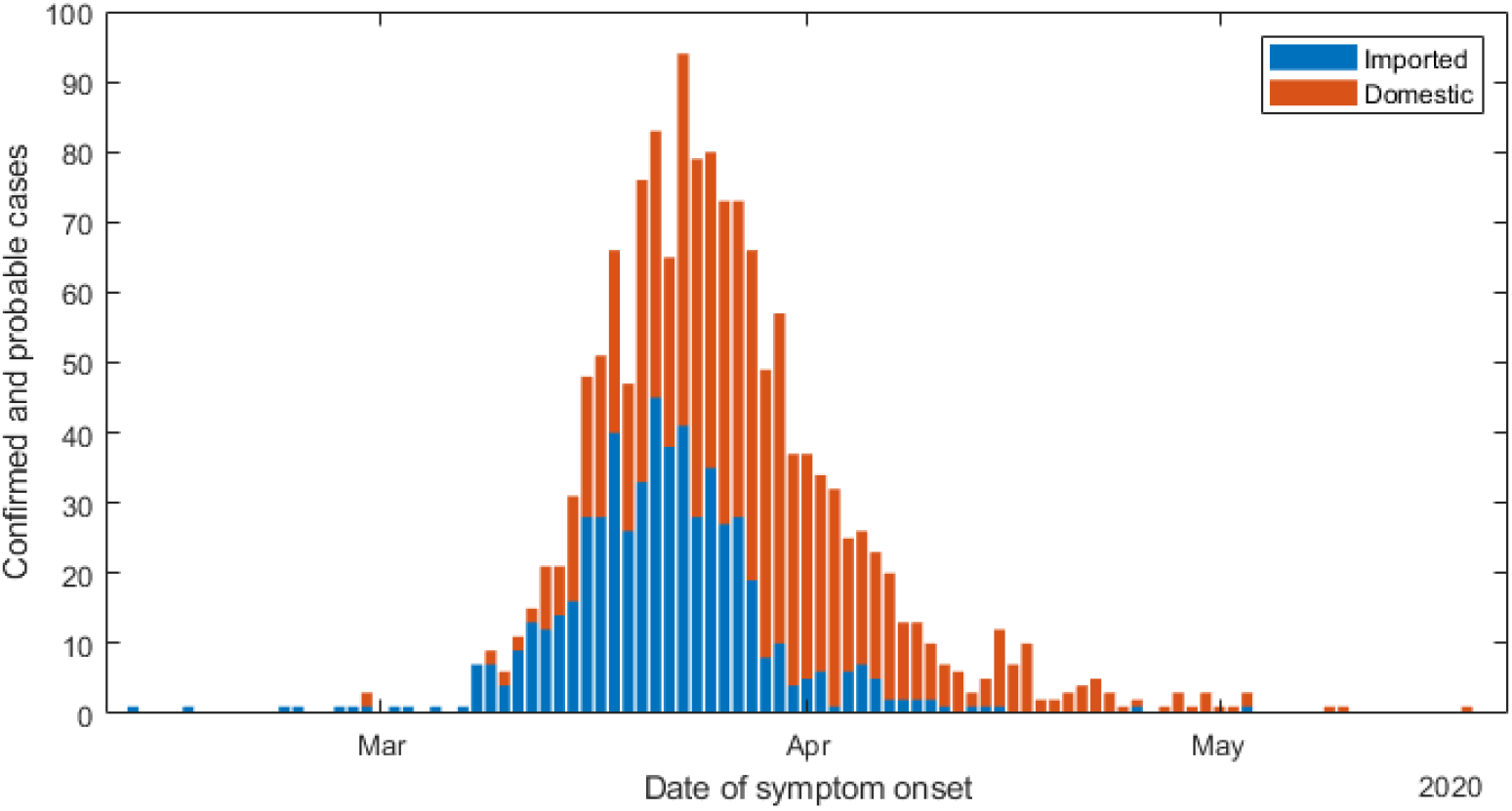
Daily new cases of COVID-19 in New Zealand from 26 February to 31 May 2020, split into cases with a recent history of international travel (blue) and those without (orange).

Progressively stricter border measures were introduced from mid-March to mid-April. From 16 March, all international arrivals to New Zealand were required to go into 14 days self-isolation. On 20 March, the border was closed to non-citizens and non-residents. From 10 April, all international arrivals were required to go into 14 day mandatory government-managed quarantine.

On 19 March, gatherings of more than 100 people were banned and, on 21 March, a four-level alert level system was introduced. Alert level 1 was effectively business as usual but with border restrictions. New Zealand entered alert level 2 on 21 March, level 3 on 23 March, and level 4 on 26 March. Alert level 4 included strict stay-at-home measures, and all non-essential business and schools were closed. The country remained at alert level 4 until 28 April, when it moved to level 3 allowing partial school openings and non-essential businesses to operate under strict non-contact guidelines. On 14 May, New Zealand moved to alert level 2 and most schools and businesses reopened but large gatherings were still banned. On 9 June, the country moved back to alert level 1 and a strict testing programme for international arrivals was established to complement the ongoing 14-day mandatory quarantine.

New Zealand also has a widespread testing regime that quickly encouraged testing of anyone with even minor symptoms. Up to 5 June (2 weeks after the last reported domestic case on 22 May), 291,994 tests were performed (58 tests per 1000 people) with a positivity rate of around 0.4% (Ministry of Health, 2020). Since mid-June, all international arrivals are tested on day three and day twelve, and are not permitted to leave their 14-day mandatory quarantine before returning a negative test result.

Mathematical models of COVID-19 transmission and control measures have estimated that the effective reproduction number *R_eff_* decreased from around 1.8 prior to alert level 4 to around 0.35 during alert level 4[1]. However, these results may depend on modelling assumptions, for example the assumed generation time distribution, the delay between infection, symptom onset and testing, and the effects of case-targeted control.

In this paper, we examine the epidemic transmission tree in New Zealand using detailed case data and contact tracing information. This is an unusual dataset because it describes a closed outbreak in which there are no ongoing transmission chains, and many cases are epidemiologically linked to a likely index case. Because not all cases have a potential index case identified and some cases have multiple potential index cases, the transmission tree is not completely or uniquely defined. We reconstruct multiple instances of the transmission tree using a Monte-Carlo technique. This approach assigns a randomly selected index case to each secondary case based on the dates of symptom onset, cluster identification, and the results of contact tracing investigations. We assume a fixed proportion of infections were undetected because they were asymptomatic, paucisymptomatic, or for some other reason.

From this suite of generated transmission trees, we calculate the individual reproduction number *R_i_* for each infected individual and explore the distribution of *R_i_* and the secondary attack rate at different points in the epidemic timeline. We explore how these quantities vary for different age groups and types of case, imported or domestic, and investigate the effect of early case isolation. We also investigate the degree of superspreading in the transmission dynamics. Although some assumptions are required to allow for missing or noisy data, our results are not model-dependent, unlike most estimates of effective reproduction number for COVID-19 [2] [3, 4]. Our sample size is larger than previous studies that have estimated the secondary attack rate (e.g. Bi, Wu (5), Ganyani, Kremer (6), Cheng, Jian (7)). This allows for more reliable estimates and for an investigation of how secondary attack rate varies with age. We also reconstruct the entire transmission tree rather than just estimating of the serial interval distribution. This provides a richer source of information about the transmission dynamics, allowing a more complete set of epidemiological parameters to be explored.

## Methods

Of the 1499 confirmed and probable cases, 575 had a recent history of overseas travel and were considered to be imported cases. The remaining 924 cases were considered to be infected within New Zealand and are referred to as domestic cases. Following contact tracing interviews, 636 of the domestic cases were each assigned up to five potential index cases. The remaining domestic cases did not have a potential index case identified. 627 of the domestic cases were linked to one of 18 significant clusters. Of the cases linked to clusters, 146 had no potential index case identified. For 1444 cases, the number of close contacts, obtained via case recall at a contact tracing interview, was recorded. The number of close contacts varied widely between individuals, e.g. 753 cases had 2 or fewer contacts and 26 cases had more than 50. 31 cases were recorded as being asymptomatic at the time of testing. All cases recorded age.

We constructed a potential instance of the transmission tree by randomly assigning a symptom onset date to each of the *i* cases using the date recorded by case recall, *D_i_*, (if available) with a random offset, *D_i_ = D_i_+ X_i_* where *X_i_* ~ *N* (*μ=* 0*,σ =* 1). For the 38 cases with no onset date recorded (31 asymptomatic, 7 symptomatic) we assigned a randomly generated onset date using the reported date and a gamma distribution fitted to the data on time from onset to reporting, Γ(*μ =* 6.7*, σ =* 5.4) days.

In addition to the 31 recorded asymptomatic cases, each generated transmission tree included additional subclinical or asymptomatic cases so that the total proportion of asymptomatic cases was *p_asymp_ =* 0.33 on average. These additional cases were randomly sampled from the recorded cases with the symptom onset date randomly jittered *D_i_ = D_i_+ X_i_* where *X_i_* ~ *N* (*μ=* 0*,σ =* 2). Model-generated asymptomatic cases had the same cluster status and potential index cases as the sampled case.

Of the domestic cases, 17 had a unique index case identified in the data. For the 288 cases with no index case, an index case was selected based on the implied serial interval, i.e. the time between symptom onset in the index case and in the secondary case. The serial interval *SI* can be expressed as

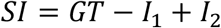

where *GT* is the generation time (time from infection of index case to infection of secondary case), and *I*_1_ and *I*_2_ are the incubation periods of the index and secondary case respectively. We assume that the generation time *GT~Weibull*(*μ =* 5.05, *σ* = 1.93) days[8] and the incubation period *I_j_~*Γ(*μ =* 5.5*, σ =* 2.28) days[9] and use this to calculate the probability density function for the serial interval *f(SI)*. Note that although the generation time must be positive, there is a positive probability of a negative serial interval.

For the 619 domestic cases with *n>* 1 potential index cases, the serial interval *SI_j_* corresponding to each potential index case *j =* 1, …*n* was calculated. Potential index case *j* was selected as the index case with probability 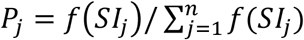. For cases with no potential index case identified, all cases in the same cluster (or the no cluster group if appropriate) were assumed to be potential index cases and the same random selection method was used. The probability *P_j_* defined above was assumed to be independent of whether the potential cases was symptomatic or asymptomatic.

Assigning an index case to each secondary case produces a single instance of the epidemic transmission tree. We then repeated this Monte Carlo process to produce *M =* 500 independent instances of the transmission tree. For each instance, we calculated the number of secondary cases resulting from each reported case (the reproduction number *R_ij_* for individual *i* in transmission tree *j*) and the associated symptom onset date, *D_ij_*.

### Incidence by age

The original case data, including clinical and asymptomatic cases, was split by imported status and time period. The reported statistic is the probability of being in each age bracket within that group and the 95% binomial confidence interval. NZ population statistics are shown as benchmarks.

### Resulting serial interval

The resulting serial interval distribution across all *M* transmission trees was fitted with a normal distribution.

### Time varying effective reproduction number

We calculated the mean reproduction number for all clinical individuals, split by imported status with onset that day (which is approximately equivalent to peak infectiousness) in transmission tree *j*. The reported statistic is the median and 95% range over the *M* transmission trees.

### Reproduction number and superspreading

We fitted a negative binomial distribution to the effective reproduction numbers in each transmission tree separately for clinical individuals of different groups (age and domestic/imported status) and with onset date *D_ij_* in different time periods for each transmission tree. This gave an estimate of the mean effective reproduction number 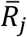 for that group of cases in the *j*^th^ transmission tree, and the associated dispersion or superspreading parameter *k_j_* [10]. We also calculated the probability of causing no secondary infections or more than 5 secondary infections for each tree. The reported statistic is the median and 95% range over the *M* transmission trees. If a negative binomial could not be fitted to a particular instance a Poisson distribution was used instead (i.e. *k = ∞*).

### Secondary attack rate

Secondary attack rates of clinical individuals were calculated for each group in each transmission tree by pooling the total number of secondary cases and the total number of contacts [11]

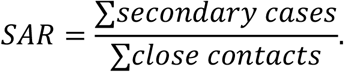

The reported statistic is the median and 95% range over the *M* transmission trees.

### Effect of early isolation

We used a general linear model to predict 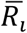, the expected reproduction number for each clinical individual across all *j* trees, accounting for days between the original recorded symptom onset and isolation, i.e. number of days for which the individual was infectious and not isolated or quarantined. Initial exploration showed no significant difference between adults and elderly so these groups were combined, children (under 10 years) were significantly different. Imported or domestic status was significant (*p* < 0.05). The number of contacts variable was also explored but this was strongly correlated with the number of infectious days which was a better predictor variable judged by the *p*-value for each coefficient (Matlab 2018b, stepwiseglm). The final model was

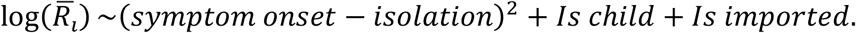

In the regression cases with 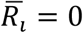 (i.e. with no secondary cases in any of the *M* instances) were offset to half the minimum value of 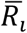 in the dataset. The results are presented graphically for an individual with the expected number of contacts.

## Results

One instance of the reconstruction of New Zealand’s nine largest clusters is shown in Figure 2. These clusters were associated with settings typical of international patterns, including a high school, a wedding, hospitality venues, aged residential care facilities, and a conference. The age distribution of cases varies across these clusters (Supplementary Figure S1). Although one of New Zealand’s largest clusters was associated with a school (Marist College cluster), more than half of the cases (55/96) in this cluster were in individuals over 20 years old. Of the 41 cases under 20, 35 were aged 10–20 years with only 6 confirmed cases in under 10s. In the two largest clusters associated with aged residential care facilities, there was as expected an overrepresentation of cases over 80 years old. However, the majority of cases in both these clusters (36/56 and 44/51 respectively) were under 65 years old, suggesting that staff and visitors are a more important driver of transmission than residents in aged residential care.

**Figure 2:**
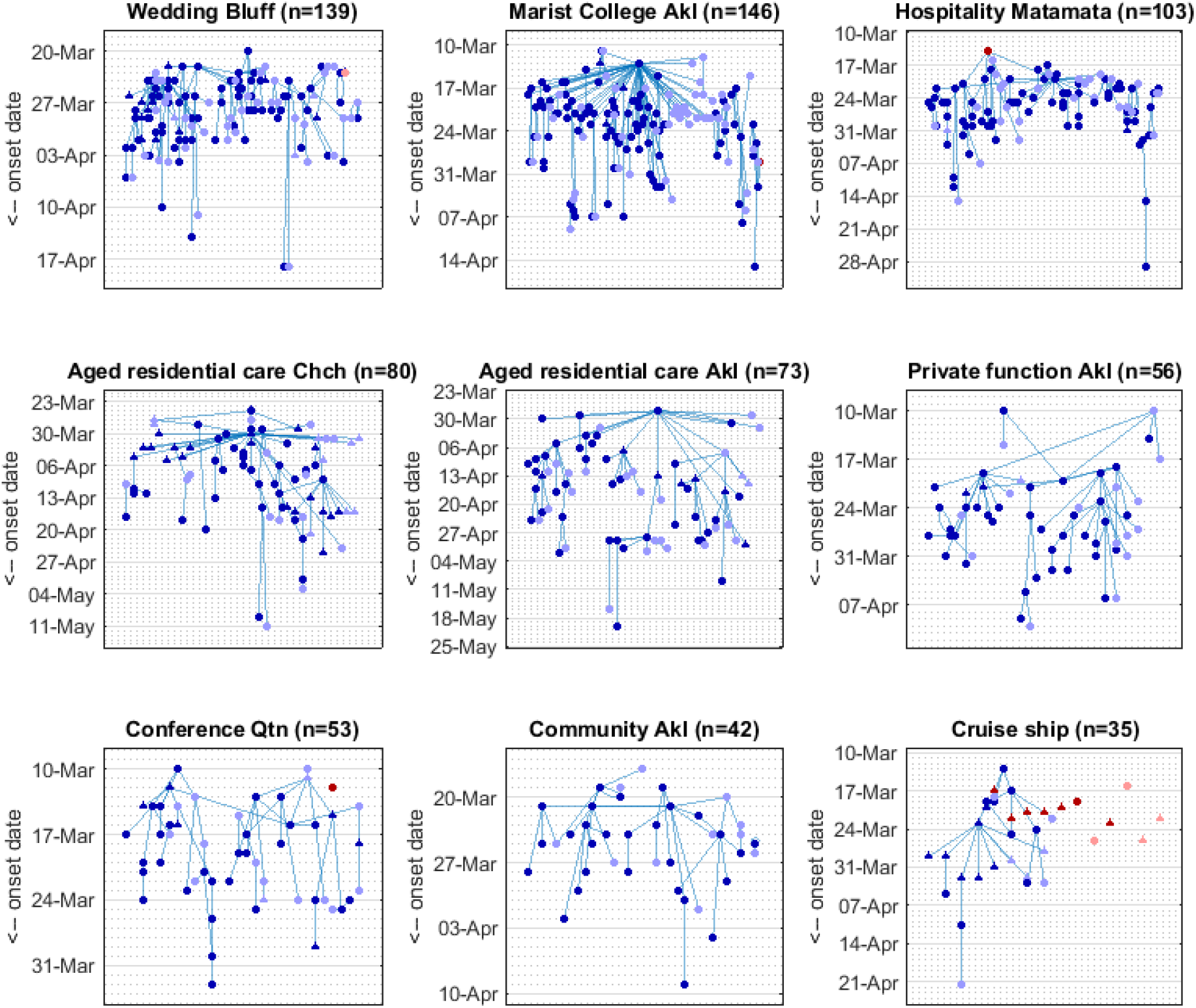
Reconstruction of New Zealand’s nine largest clusters. Each panel shows one instance of the Monte Carlo method for reconstructing the epidemic transmission tree with cases plotted by date of symptom onset. Red = imported case; blue = domestic case; dark colours = symptomatic; light colours = asymptomatic; squares = age under 10 years; circles = age 10-65 years; triangles = age over 65 years. The title of each panel describes the setting and location (Akl = Auckland; Chch = Christchurch; Qtn = Queenstown) associated with each cluster; n is the total number of cases in the reconstructed tree including actual cases and simulated (missing) asymptomatic cases.

### Incidence by age

We investigated COVID-19 incidence by age by splitting the clinical cases into three age groups: under 10 years, 0-65 years, and over 65 years. We consider domestic and imported cases separately and further split the data by significant time periods: before and after alert level 4 was introduced (26 March) for the domestic cases; and during three distinct periods of border control for imported cases: no restrictions (before 16 March), self-isolation (16 March to 9 April) and government-mandated quarantine (after 9 April).

Children under 10 are significantly under-represented in the case data relative to the New Zealand population demographic of 12.7% (Figure 3). The domestic cases (Fig 3 A,B) contain relatively more children under 10 than the imported cases and these infections predominantly occurred during alert level 4 (2.6% of cases were in children before 26 March, 4.4% on or after 26 March). During alert level 4, schools and day care centres were closed so children are likely to have spent more time in family settings where attack rates are higher [5] resulting in a higher incidence for this group. Of the 576 imported cases (Fig 3 C-E), only 2 were in children under 10 years old. Both these cases were connected with an imported adult case with the same travel history and symptom onset 4 to 7 days prior.

**Figure 3:**
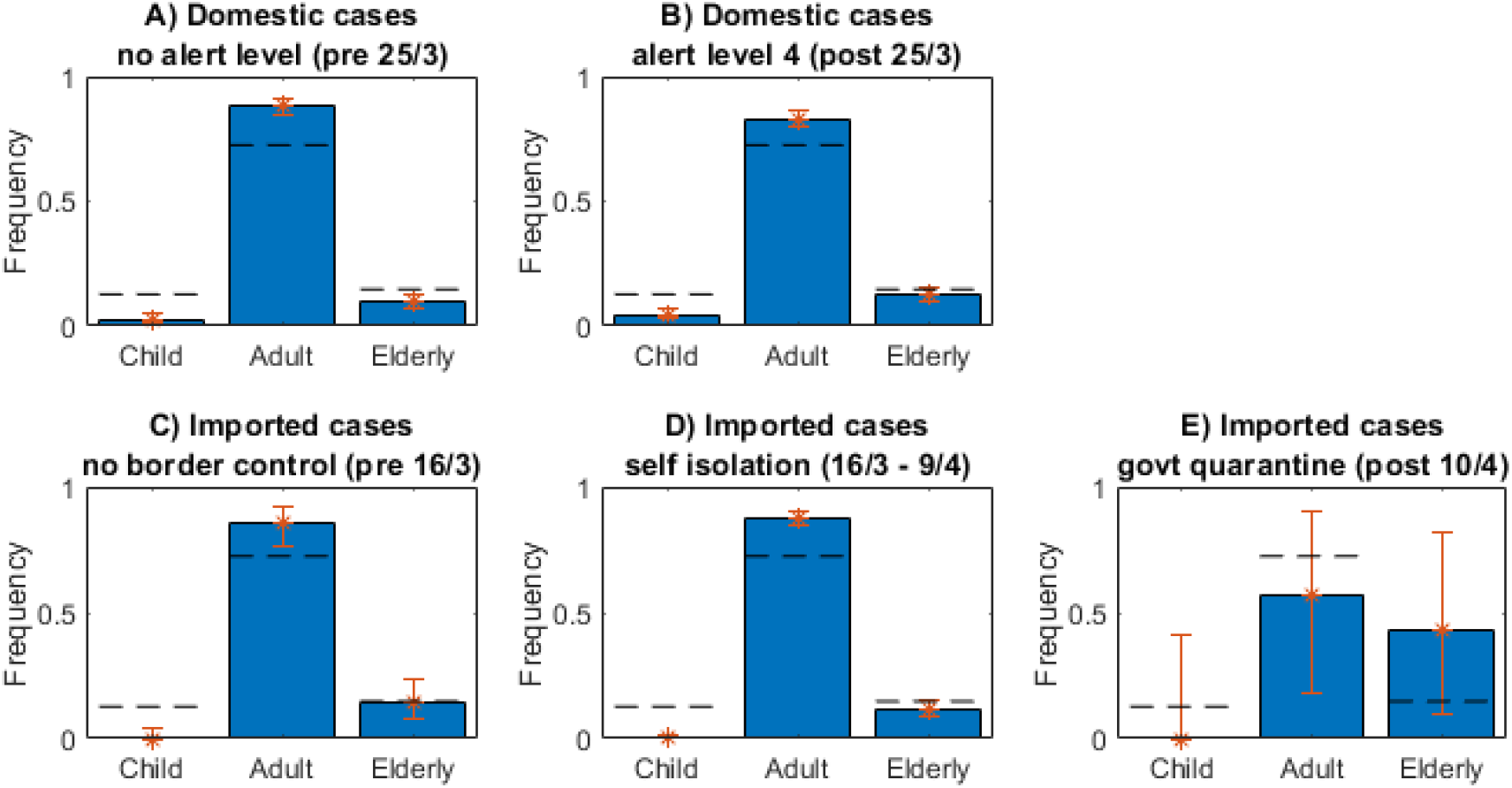
Across all categories of case children are under-represented. Frequency of cases for the different types of case aged: children under 10 years; adults 10-65 years; elderly over 65 years. Dashed lines show New Zealand population. Error bars show the binomial 95% confidence interval.

### Serial interval distribution

The distribution of serial intervals (Figure 4) resulting from the Monte Carlo reconstruction contained a substantial number of outliers. It is likely these correspond to cases that were infected by an undetected case, or the index case was wrongly assigned either in the data or in the reconstructed tree. Including all serial intervals from the reconstructed tree, the best fit normal distribution had mean 5.0 days and standard deviation 5.7 days. Excluding outliers (serial intervals longer than 20 days) resulted in mean 4.4 days and standard deviation 4.7 days. This is comparable with previous estimates of the mean serial interval [12, 13]. When allowing for negative serial intervals in their Monte Carlo reconstruction method, Ganyani, Kremer (6) found the serial interval to be 3.86 ± 4.76 days (mean ± std. dev.) for Singapore and 2.90 ± 4.88 days for Tianjin, China. These estimates for the mean serial interval are shorter than ours, but the finding that a significant proportion of serial intervals were negative is consistent. The data we analysed and that of Ganyani, Kremer (6) were collected in the presence of case isolation and contact tracing. It is likely these interventions will reduce the mean serial interval, to varying extents, by suppressing transmission late in the infectious period. Our results reflect the serial interval distribution in the presence of these interventions in the New Zealand context.

**Figure 4.**
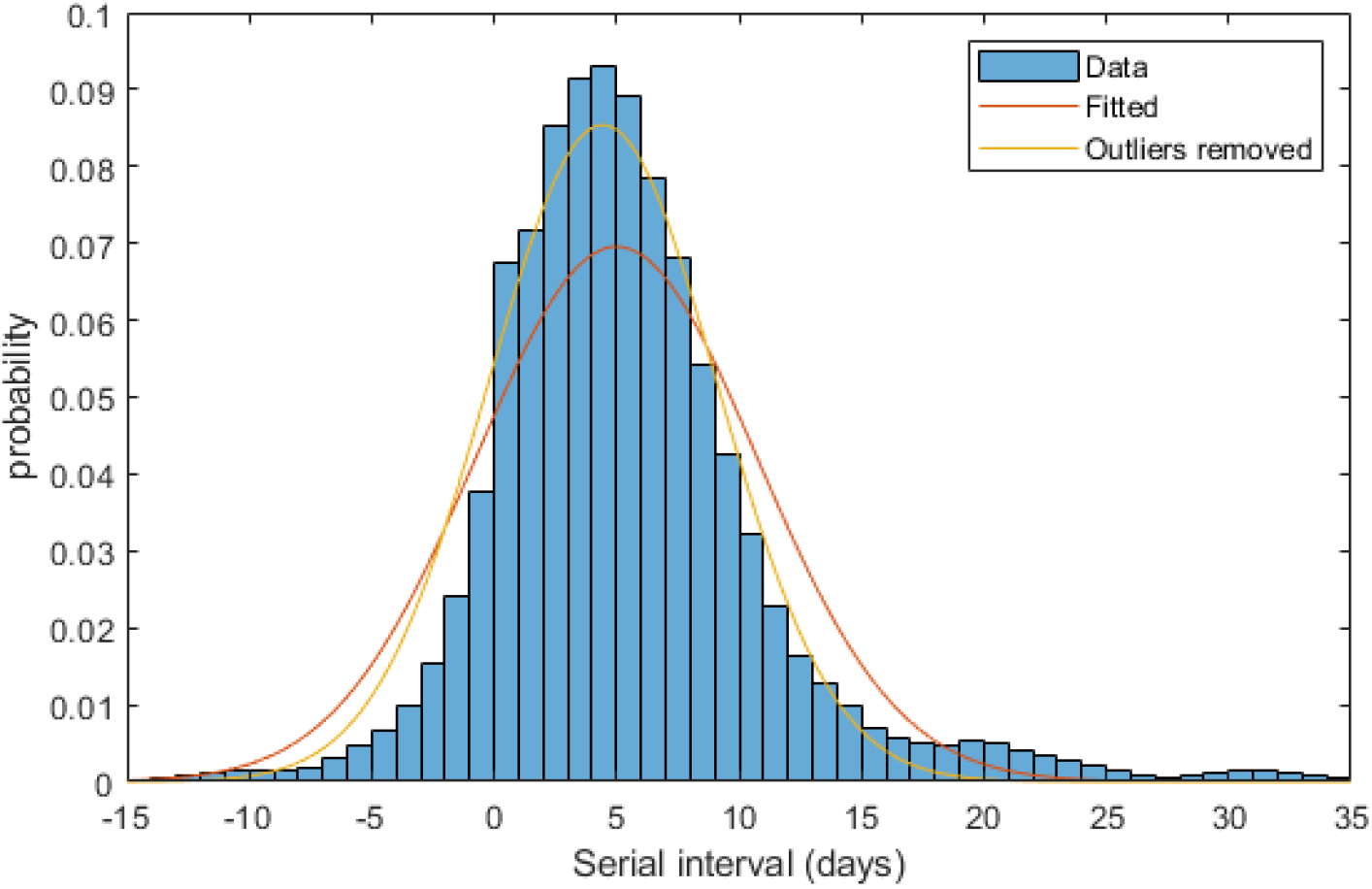
Serial interval distribution with fitted distributions. Red curve shows the normal distribution fitted to all data (*~N*(*μ* = 5.0, *σ* = 5.7)); yellow curve shows the normal distribution fitted to data excluding serial intervals longer than 20 days (*~N*(*μ =* 4.4*, σ =* 4.7)).

### Time-varying effective reproduction number

For domestic cases, the average number of secondary cases infected by a clinical case before New Zealand entered alert level 4 on 26 March was *R_eff_* = 1.47 (382 index cases) (Figure 5A). During and after alert level 4 (26 March onwards), this dropped to *R_eff_* = 0.69 (521 index cases). In the period after alert level 4, the vast majority of cases were detected as part of contact tracing efforts and were isolated swiftly, often before symptom onset. The reproduction number for domestic cases showed two small peaks during the alert level 4 period. The first coincided with Easter weekend (10-13 April). The second coincides with a long weekend (25-27 April) immediately preceding the pre-announced easing of restrictions on 28 April.

**Figure 5:**
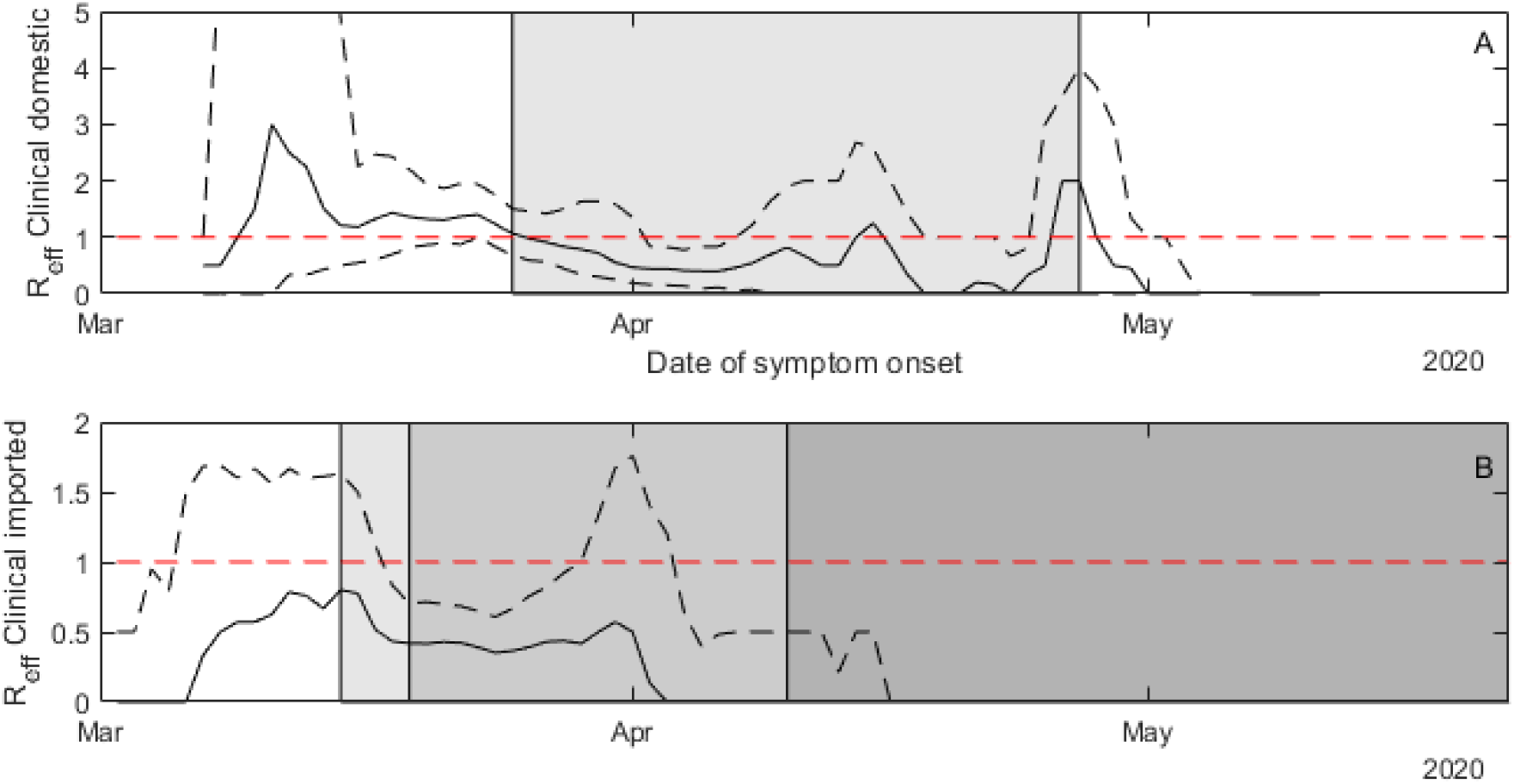
Increases in alert level and improved border measures resulted in decreases in secondary transmissions. Expected value of *R_eff_* each day for clinical domestic cases (A) and imported cases (B). Solid line show the median from 500 instances of the reconstructed transmission tree, dashed lines are the 95% range. Imported cases had a lower effective reproduction number than domestic cases. The date of secondary case exposure was assigned to be the date of symptom onset in the index case. Note that 10-13 April was the long Easter weekend and 27 April was a public holiday in New Zealand. The timing of the end of alert level four was announced on 22 April. Shading in (A) shows period of alert level 4 (26 March to 27 April). Shading in (B) shows progressively stricter border restrictions introduced 16 March, 20 March and 10 April.

For imported cases (Figure 5B), the number of secondary infections per case was much lower throughout the outbreak. Before self-isolation for international arrivals was introduced on 16 March, *R_eff_ =* 0.70 (119 index cases). While self-isolation was in place from 17 March to 9 April, *R_eff_* = 0*A*2 (439 index cases). After mandatory government-managed quarantine was introduced on 10 April, *R_eff_* = 0.08 (7 cases).

### Effect of age on reproduction number and superspreading

Before the move to alert level 4, over half of all domestic cases resulted in at least one secondary case (Figure 5A). The proportion of cases resulting in at least one secondary case was similar for all age groups (Table 1). The expected number of secondary cases was *R_eff_* = 0.87 for the under 10 years age group, *R_eff_* = 1.49 for the 10 to 65 years age group, and = 1.51 for the over 65 years age group. Although children under 10 were equally likely to infect at least one person, adults tended to infect more people than children under 10 did. Cases among adults and older people also had a significant chance of being a superspreader (which we defined as infecting more than 5 people): 6% in the 10 to 65 year age group, and 7% in the over 65 age group.

**Table 1:**
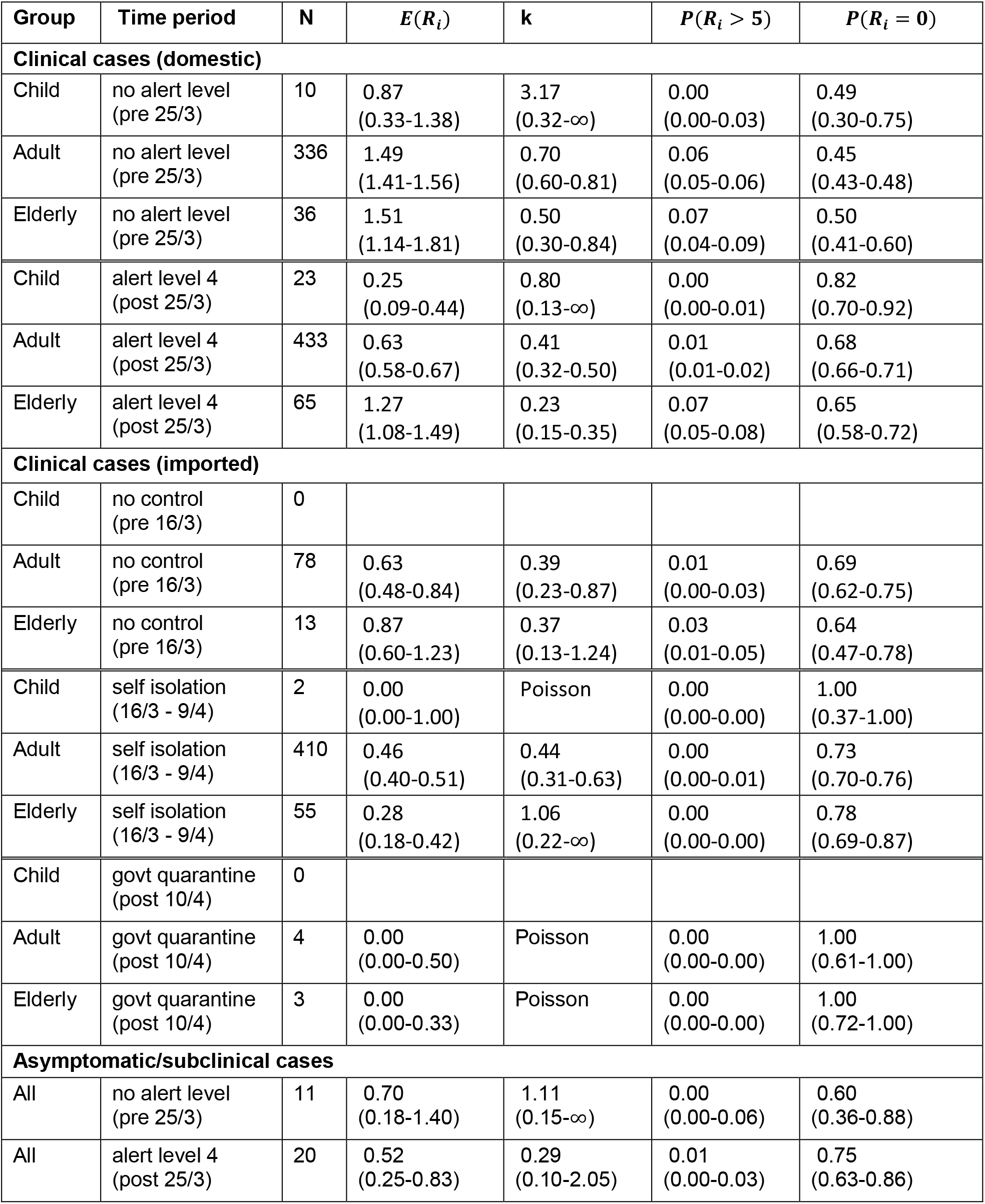
Summary statistics for the New Zealand epidemic by age and type of case. Columns show: the number of cases in each group (N); the expected number of secondary infections caused by an individual in this group (*E*(*R_i_*)); the dispersion coefficient of a fitted negative binomial distribution (k); the probability of an individual infecting more than 5 people (***p***(***R_i_*** *>* **5**)) and the probability of an individual not infecting anyone (***p***(***R_i_*** = **0**)) according to the fitted distribution. Numbers show the median (95% range) across 500 instances of reconstructed transmission tree.

During alert level 4 (Fig. 5B, Table 1), *R_eff_* dropped below 1 for the under 10 and the 10 to 65 age groups, but stayed above 1 in over 65 group. This may be due to the over-representation of cases in aged care facilities in the later stages of the epidemic, where it may be more difficult to limit interactions. The proportion of superspreaders decreased in adults during alert level 4 but not in the elderly. There were no recorded superspreaders under 10 years old at any stage in the epidemic.

Overall, there were 29 superspreaders, of which 21 had symptom onset before alert level 4 began, and the remaining 8 had symptom onset during alert level 4. Of these, the 6 most recent events were all related to clusters of cases at aged care facilities.

As seen previously, imported cases overall had a lower *R_eff_* than domestic cases (Figure 6C-E). Before the introduction of any border measures, many of these cases would have been tourists and other visitors who are less likely to have the close community contacts that local residents have. Throughout the outbreak, imported cases in the over 65 years age group had a higher *R_eff_* than those aged 10 – 65. Imported cases in children, of which there were only 2 (<1%), had the lowest likelihood of being an index case even accounting for both these cases happening during the mandatory quarantine period. There were 5 superspreaders amongst the imported cases, all of which occurred during the self-isolation period between 9 and 19 March.

**Figure 6:**
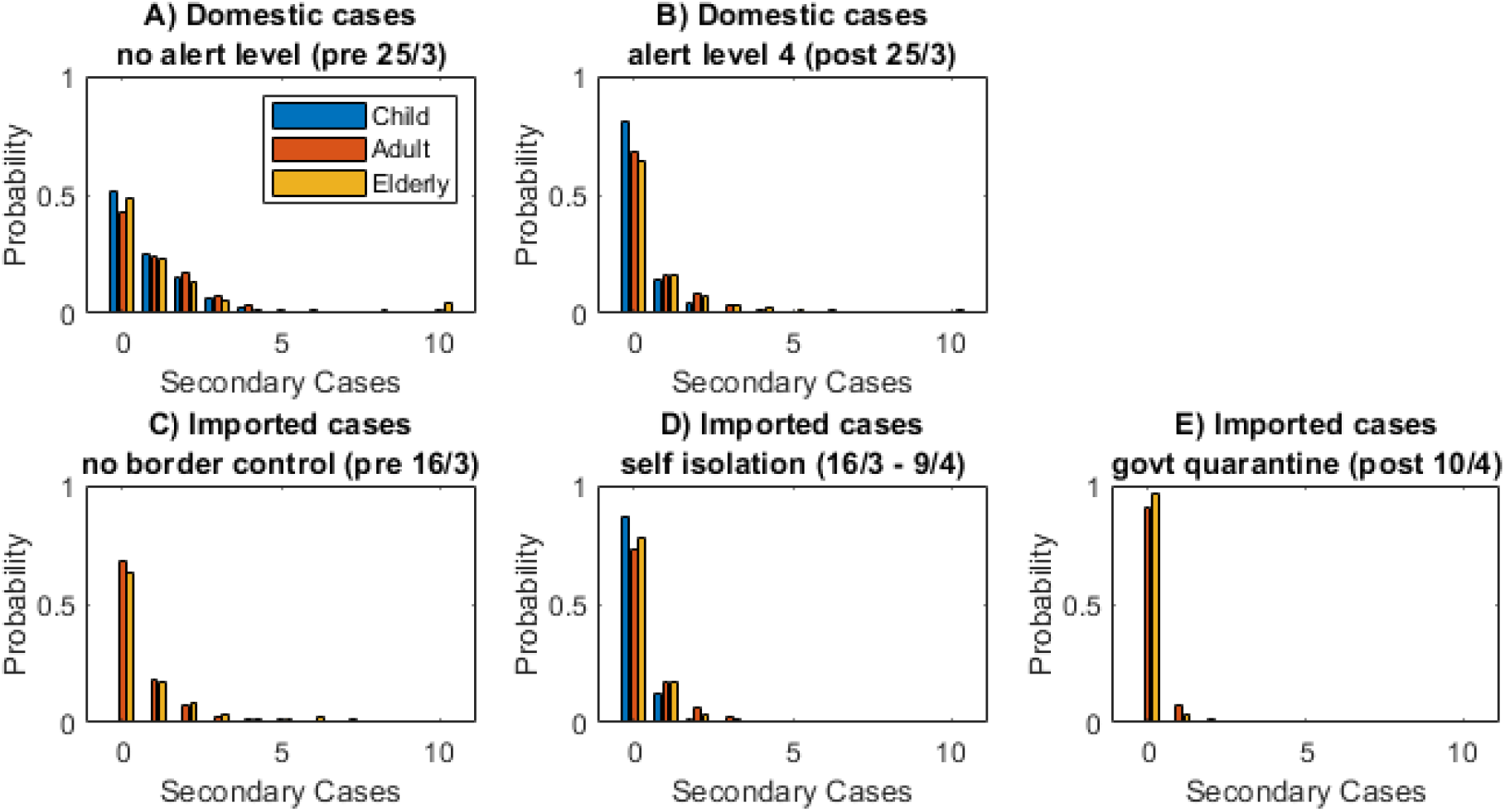
Children under 10 tended to infect fewer people and were less likely to be superspreaders. Distribution of individual reproduction numbers for domestic and imported cases in each age group: under 10 years (blue), 10 to 65 years (red), over 65 years (yellow).

The degree of superspreading in the transmission dynamics can be quantified by the variance in the distribution of individual reproduction numbers 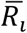. This can be measured by the dispersion parameter *k* of a negative binomial distribution fitted to the data [10, 14]. The smaller the value of *k*, the more heavy-tailed the distribution of secondary cases, and therefore the more superspreading is driving the transmission process. The results show that older age groups tend to be have more superspreading than younger ones (Table 1). This could be influenced partly by the number of cases transmitted in aged residential care facilities. Among the 10-65 and over 65 year age groups, the estimated values of *k* range from 0.27 to 0.70. This means that 20% of cases are responsible for between 65% and 85% of transmission.

### Secondary attack rate

In all case groups, children under 10 years old had a lower secondary attack rate than the older age groups (Figure 7). For domestic cases in the under 10 and the 10–65 age groups, there was very little difference in secondary attack rate before and during alert level 4, despite contacts being largely confined to the home setting which has a higher attack rate [5]. The secondary attack rate for elderly people did show a large increase during alert level 4. This likely reflects the number of cases that occurred in aged care facilities rather than private households.

**Figure 7:**
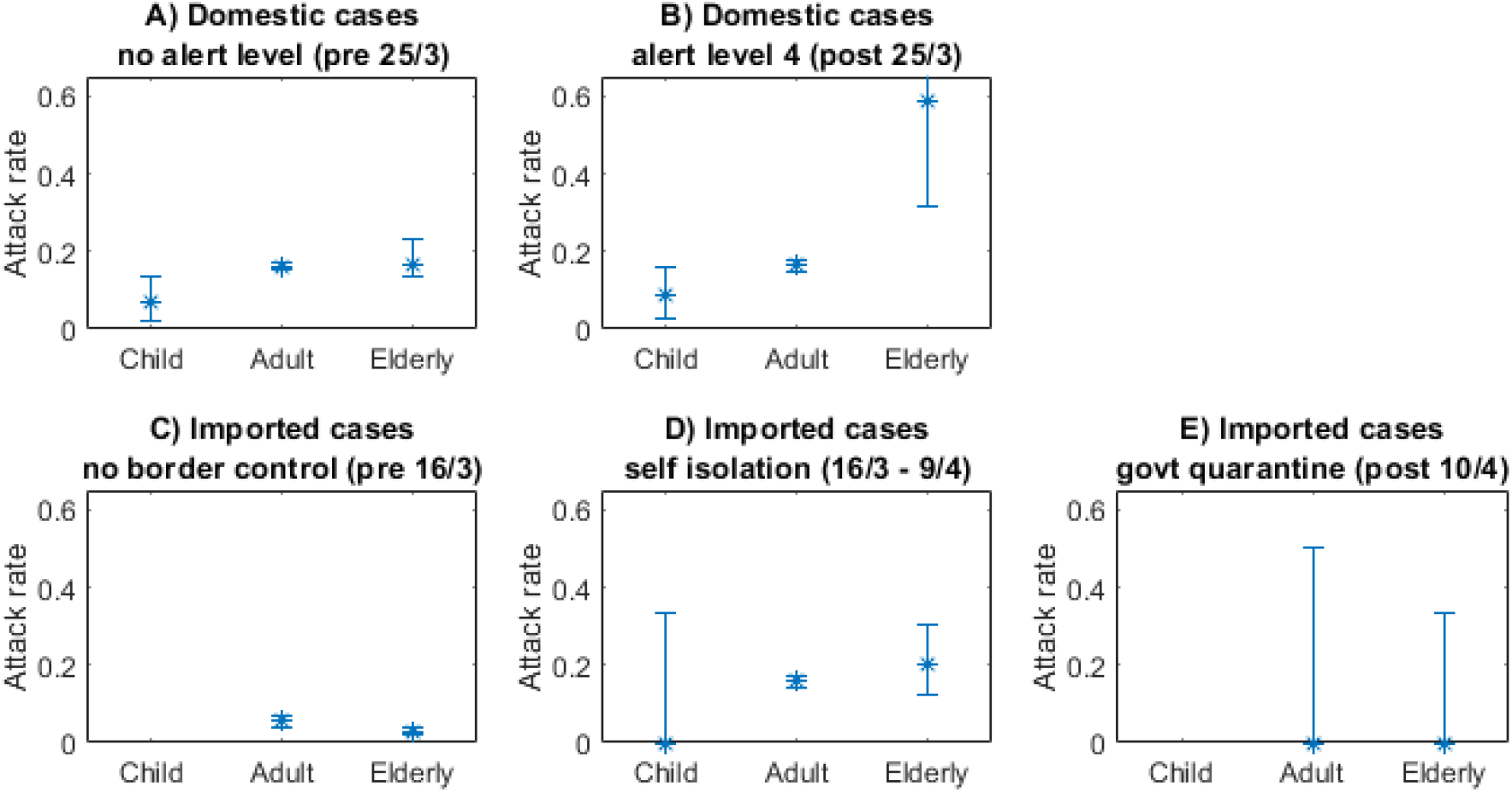
Children under 10 had a lower secondary attack rate across all case groups. Secondary attack rate for each group by age.

Overall, the secondary attack rate for imported cases was much lower than for domestic cases, particularly during the period before introduction of border measures. This is likely due to this group including a high proportion of visiting tourists for whom most contacts are in a casual rather than home or work setting. The secondary attack rate for imported cases increased after the self-isolation measures were introduced, possibly reflecting the environment and make-up of these groups. However, the secondary attack rate dropped significantly during the mandatory quarantine period.

The lack of a clear effect of control measures on secondary attack rates suggests that reduction in the reproduction number was achieved primarily by reducing the number of close contacts rather than by reducing the likelihood of transmission when close contacts did occur.

### Effect of case isolation

Figure 8 shows the general linear model for the relationship between the individual reproduction number 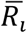 and the time between symptom onset and isolation. This shows that cases that were isolated earlier, relative to symptom onset, tended to infect fewer people. Accounting for overseas status and number of infectious days before isolation or quarantine children infect, on average, 62% the individuals that adults and the elderly infect (*p* = 0.09). Imported cases infect 70% of the number of secondary cases in comparison to domestic cases (*p* = 10^−5^).

**Figure 8:**
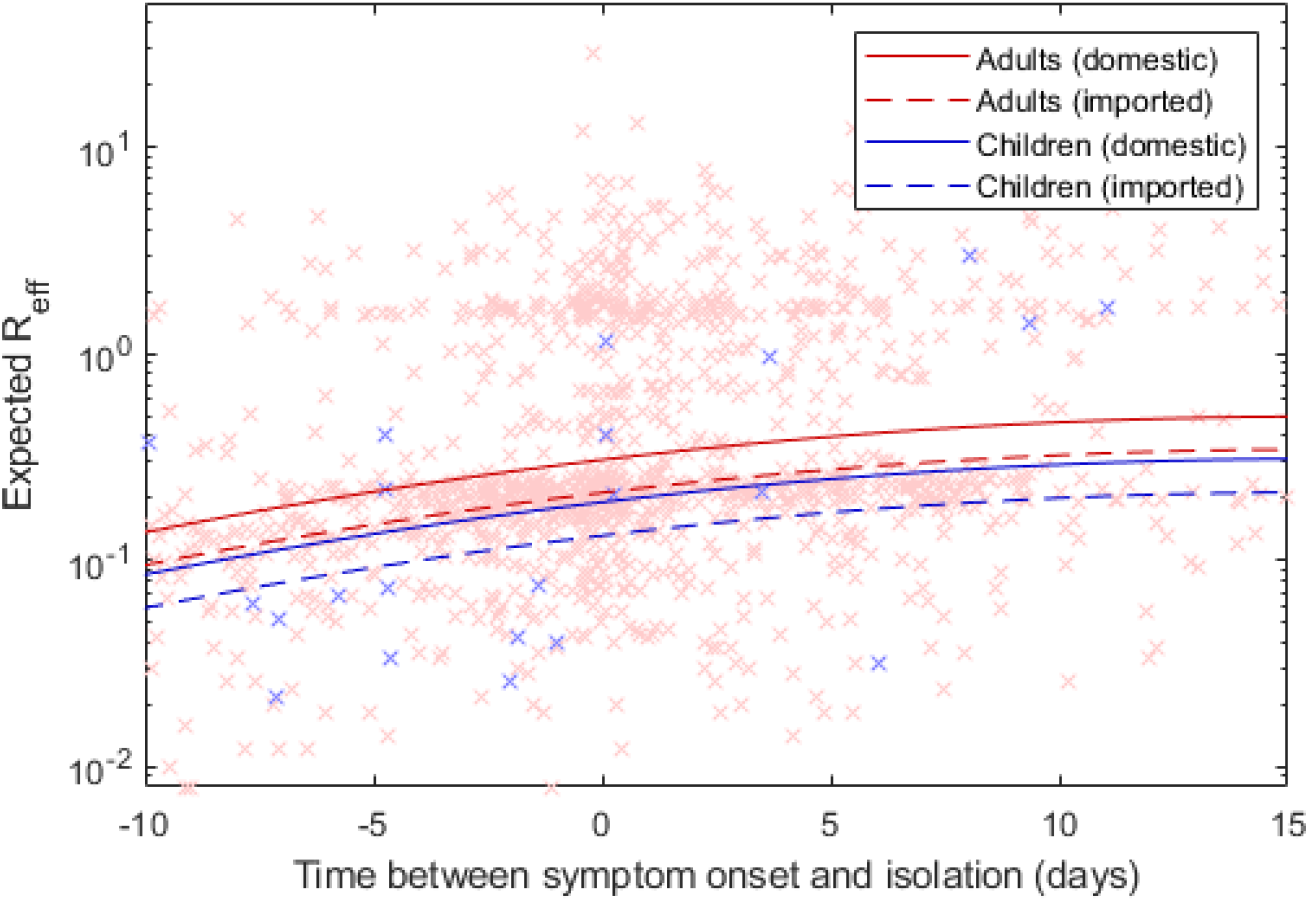
Early isolation substantially decreases secondary transmission and children and imported cases have a significantly lower transmission rate compared to adults and domestic cases. The expected reproduction for each individual across all 500 trees (red crosses adults and elderly, blue crosses under 10 year olds) against number of infectious days measured as time from symptom onset to isolation. Lines are the output of a regression model for each class of individual.

### Asymptomatic cases

Of the 31 cases (21 domestic, 10 imported) recorded as asymptomatic, 30 were in adults and older people. Before the move to alert level 4, these cases had *R_eff_* of approximately 50% of the adult domestic clinical cases (Table 1). During alert level 4, this dropped slightly further but the drop was not as significant as or the clinical cases during this time.

## Discussion

We analysed the transmission dynamics of COVID-19 using a comprehensive dataset for a completed outbreak. Our analysis investigated the role of age and type of case (imported or domestic) on reproduction number, secondary attack rate, likelihood of superspreading. We also investigated the effect of various control measures, including case isolation, population-wide restrictions, and self-isolation or quarantine of international arrivals.

We found that children under 10 years contribute less to the spread of COVID-19 than other age groups. Children under 10 infected fewer people on average, had a lower secondary attack rate, and were less likely to be superspreaders. This is consistent with existing evidence on the role of children in COVID-19 transmission [15, 16].

The finding that superspreading is a significant contributor to transmission is consistent with existing empirical and modelling studies [17] [18] [19]. Our results show that among adults 20% of cases are responsible for between 65% and 85% of transmission. This suggests that interventions targeting superspreaders or superspreading events may be particularly effective in reducing the spread of COVID-19. These may include restrictions on gathering size, particularly in closed environments or crowded spaces [17].

In the dataset we used to reconstruct epidemic transmission tree, the index case is either missing or not uniquely identified in some cases. It is also likely there were asymptomatic or paucisymptomatic cases that were undetected. We therefore used Monte Carlo methods to reconstruct missing cases and missing transmission routes (nodes and links respectively in the epidemic tree). It is possible that phylodynamic data on the genetic sequence of the virus recovered from positive test results could be used to reduce uncertainty in this step and improve the accuracy of the reconstructed tree.

## Data Availability

For privacy reasons the data is not publicly available. Contact the New Zealand Ministry of Health for research related data requests.

## Acknowledgements

The authors are grateful to Annette Nesdale of the Hutt Valley District Health Board for discussions about the data analysed in this study. We acknowledge the support of StatsNZ, ESR, and the Ministry of Health in supplying data in support of this work. This work was funded by the Ministry of Business, Innovation and Employment and Te Punaha Matatini, New Zealand’s Centre of Research Excellence in complex systems.

## Supplementary Material

**Supplementary Figure S1.**
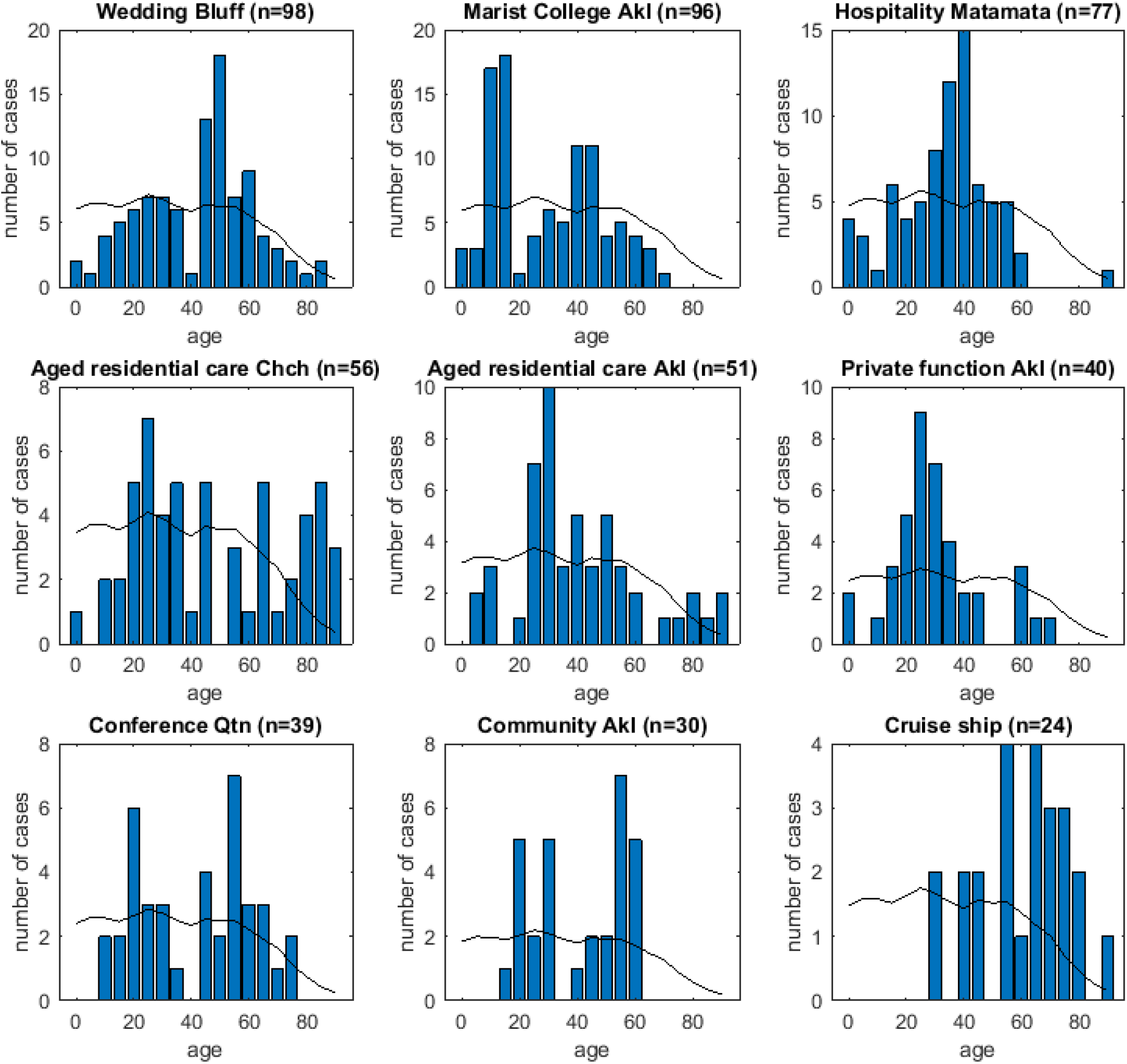
Age distribution of cases in New Zealand’s nine largest clusters. The black line shows the age distribution of the New Zealand population (number of cases that would be expected if the cluster followed the same age distribution as the New Zealand population). The title of each panel describes the setting and location (Akl = Auckland; Chch = Christchurch; Qtn = Queenstown) associated with each cluster; n is the total number of recorded cases in the cluster.

